# Genomic epidemiology of SARS-CoV-2 in Sudan: A retrospective analysis 2020-2022

**DOI:** 10.1101/2023.11.27.23299044

**Authors:** Maureen W. Mburu, Arnold W. Lambisia, Musab Elnegoumi, Eman O. M. Nour, Osama M. M. Khair, Elamin Abualas, Rehab M Elhassan, Muatsim A. M. Adam, Khadija Said Mohammed, John M. Morobe, Leonard Ndwiga, Edidah O. Moraa, Timothy O. Makori, Nickson Murunga, Esther Nyadzua Katama, Dorcas W. Waruguru, Sarah Mwangi, Sofonias K. Tessema, Mawahib H. E. Adam, Iman Mahmoud, Hanan SalmanBabikir Mohamed, Yousif A. A. Mustafa, Mohamedtaha M. A. Mohamed, Rasheeda H.A. Ahmed, Nuha Y.I. Mohamed, Rahma H. Ali, Rasha S.M. Ebraheem, Huda H.H. Ahmed, Hamadelniel E. Abdalla, Izdihar Mukhtar, Abdalkhalig E. M. W. Alla, Omer H. A. Elsheikh, Abdualmoniem O. Musa, Mahmoud Taha Faki, Gada Mustfa Ahmed, Thoyba E. R. Ahmed, Mohammed A. O. Abdalla, Mujtaba O. K. Mohamed, Rayan A. A. Ahmed, Samah M. M. AwadElkarim, Basamat A. E. M. Ahmed, Zainab A. I. Shumo, Eiman N. A. Ahmed, Samia S. A. Rushwan, Phillip Bejon, D. James Nokes, Lynette Isabella Ochola-Oyier, Charles N. Agoti, George Githinji, Shahinaz A. Bedri

## Abstract

SARS-CoV-2 was first detected in Sudan on 13^th^ March 2020. Here, we describe the genomic epidemiology of SARS-CoV-2 in Sudan between May 2020 and April 2022 to understand the introduction and transmission of SARS-CoV-2 variants in the country. A total of 667 SARS-CoV-2 positive samples were successfully sequenced using the nCoV-19 Artic protocol on the Oxford Nanopore Technology (≥70% genome completeness). The genomes were compared with a select contemporaneous global dataset to determine genetic relatedness and estimate import/export events. The genomes were classified into 37 Pango lineages within the **ancestral strain** (107 isolates across 13 Pango lineages), **Eta variant of interest (VOI)** (78 isolates in 1 lineage), **Alpha variant of concern (VOC)** (10 isolates in 2 lineages), **Beta VOC** (26 isolates in 1 lineage), **Delta VOC** (171 isolates across 8 lineages) and **Omicron VOC** (242 isolates across 12 lineages). We estimated a total of 144 introductions of the observed variants from different countries across the globe. Multiple introductions of the Eta VOI, Beta VOC and Omicron VOC were observed in Sudan mainly from Europe and Africa. These findings suggest a need for continuous genomic surveillance of SARS-CoV-2 to monitor their introduction and spread consequently inform public health measures to combat SARS-CoV-2 transmission.

## INTRODUCTION

The first confirmed SARS-CoV-2 case in Sudan was detected on 13^th^ March 2020 and by end of April 2022 the country had confirmed a total of 62,131 cases and 4,933 COVID-19 related deaths [1]. A population-based study conducted between 1^st^ March to 10^th^ April 2021 in Omdurman city in Sudan estimated a seroprevalence of 54.6% with higher SARS-CoV-2 seropositivity among older people (>50 years)[2].

The first major SARS-CoV-2 wave of infections was observed between 7^th^ April and 22^nd^ September 2020 [1]. The government introduced various countermeasures such as closure of schools, airports and borders, restriction of movement across localities, dusk to dawn curfews and banning of religious and social gatherings in order to curb the spread of the virus, but these measures were relaxed from September 2020 [3]. Due to challenges in contact tracing earlier in the pandemic in Sudan, the SARS-COV-2 cases persisted and resulted in the second surge of infections between November 2020 and January 2021 [2] [3]. Notably, political instability in the country affected tracking and containment of SARS-CoV-2 infections. The third major wave was observed between December 2021 and February 2022 [1].

The major waves of SARS-CoV-2 cases observed in Sudan coincided with the infection waves observed in other African countries and these have been attributed to import/export events between different countries globally [4]. Sudan is bordered by seven countries namely, Eritrea, Ethiopia, South Sudan, Central African Republic, Chad, Libya, and Egypt with high movement across the borders and major cities lying either along the Red Sea or Nile River. Previously, transmission of viral infectious diseases such yellow fever and recurrent arbovirus outbreaks have attributed to rural-urban migration within the country[3]. However, the transmission dynamics of SARS-CoV-2 variants in Sudan and across its borders during the pandemic to date has not been previously described.

Genomic surveillance has been instrumental in monitoring and understanding SARS-CoV-2 variants evolution and spread in many countries globally [5]–[8]. This study aims to describe SARS-CoV-2 variants and transmission dynamics in Sudan using 667 whole genome sequences obtained from PCR-confirmed cases collected between May 2020 and April 2022.

## METHODS

### Ethical approval

The genomic surveillance protocol used at Kenya Medical Research Institute (KEMRI) Wellcome Trust (KWTRP) was reviewed and approved by the Scientific and Ethics Review Committee (SERU) in KEMRI (SERU #4035). The SARS-CoV-2 positive samples were part of the public health emergency response thus individual participant consent was waived.

### SARS-CoV-2 sampling

A total of 667 SARS-CoV-2 positive samples were sent to KWTRP-Kenya, ILRI-Kenya and South Africa for whole genome sequencing. The dates of collection spanned from 20^th^ May 2020 to 18^th^ April 2022.

### Viral RNA extraction and cDNA synthesis

SARS-CoV-2 samples were shipped from the National Public Health Laboratory – Sudan to KWTRP-Kenya (n=405). The samples shipped to KWTRP-Kenya were extracted using QIAamp Viral RNA Mini kit (52906, Qiagen, Hilden, Germany) following the manufacturer’s instructions. SARS-CoV-2 positivity screening was done using various quantitative PCR kits. Samples with a cycle threshold (Ct) ≤ 34 were selected for cDNA synthesis. 8µl of RNA was reverse transcribed using 2 µl LunaScript® RT SuperMix Kit.

### Whole genome amplification, library preparation and sequencing at KWTRP-Kenya

The SARS-CoV-2 genome was amplified using Q5® Hot Start High Fidelity 2X Master Mix (NEB, US) and the modified V4 ARTIC primers [9]. The amplicons were cleaned using 1x AMPure XP beads (Beckman Coulter, US) as per the manufacturer’s protocol, eluted in nuclease free water and the eluent quantified using Qubit dsDNA HS Assay Kit (ThermoFisher Scientific, US). Library preparation was performed as previously described [9], [10].

End-prep was done using 50ng of PCR amplicons according to the ARTIC nCoV-2019 sequencing protocol v3 (LoCost) [11] using the NEBNext Ultra II End repair/dA-tailing Kit (NEB, US). Barcode ligation was done using the native barcoding expansion (EXP-NBD196) kit (Oxford Nanopore Technology, UK) and NEBNext Blunt/TA Ligase Master Mix (NEB, US). Thereafter, adaptor ligation was done using NEBNext Quick Ligation Module reagents (NEB, US), Adaptor Mix II (ONT, UK) and 100ng of the barcoded sample following the manufacturers instruction. The final loading concentration was normalised to 15ng and loaded on a SpotON R9 flow cell and sequenced on a GridION device [12].

### SARS-CoV-2 genome assembly

Basecalling and demultiplexing were executed simultaneously during sequencing using Guppy version 4.0.5 (https://artic.network/ncov-2019/ncov2019-bioinformatics-sop.html) with a minimum Q score of 1. Consensus SARS-CoV-2 genomes were generated using the ARTIC bioinformatics protocol (https://artic.network/ncov-2019/ncov2019-bioinformatics-sop.html). Briefly, the reads were filtered based on length (300-700bp) and mapped to the reference genome (GenBank accession MN908947.3) using Minimap2 (https://github.com/lh3/minimap2). Fast5 reads were used to polish the consensus genomes using Nanopolish (https://github.com/jts/nanopolish) and consensus genomes generated with a read depth of 20X.

### Additional Sudan data

Additional sequences from Sudan generated at ILRI-Kenya (n= 199) and South Africa (n=204) available in the Global Initiative on Sharing All Influenza Data (GISAID) were also included in the subsequent analysis [13].

### Lineage assignment, phylogenetics and mugration analysis

The consensus genomes were assigned lineages using Pangolin version 4.0.6 [13]. Clade assignment and mutation profiling was done using NextClade v1.13.2 [14].

All sequences originating from Sudan were analysed against a global dataset of 13,122 SARS-CoV-2 genomes to ensure that global representation of sequences. The selected global subset had ≥95% genome coverage and were selected evenly from across the Pango lineage, time (month of collection) and continents. The genomes were collected from 1^st^ September 2020 to 19^th^ January 2022. All global reference genome sequences and associated metadata in this dataset are published in GISAID’s EpiCoV database https://doi.org/10.55876/gis8.220815ga.

The sequences from Sudan and the global dataset were aligned using Nextalign version 1.4.1 (https://github.com/neherlab/nextalign) against the SARS-CoV-2 reference genome. A maximum likelihood (ML) tree was generated using IQTREE version 2.1.3 (http://www.iqtree.org/) and a branch support was assessed using 1000 bootstrap iterations. The ML trees were converted to time resolved phylogenetic trees using TreeTime (https://github.com/neherlab/treetime).

A Python script was used to estimate SARS-CoV-2 import and export events in Sudan based on the origin of sequence and time resolved tree’s tips and internal nodes. The import/export events were visualized using R packages alluvial and tidyverse [15]

## RESULTS

### Baseline demographic characteristics

A total of 667 SARS-CoV-2 samples were collected from 11 different locations in Sudan and a majority of these samples were from Khartoum (n=390, 58.5%), Kassala (n=180, 27%) and Port Sudan (n=66, 9.9%). Among this population, the median age was 40 years. Out of the 667 cases, 237 (35.5%) were presented with symptoms and only 4 (0.6%) had known international travel history. Metadata for variables such as vaccination status, travel history, sex, age, and reason for testing was lacking for >10% of the cases (**Supplementary table 1**).

### SARS-CoV-2 variants surveillance in Sudan

Genomic surveillance of SARS-CoV-2 started in May 2020 in Sudan and the 667 genomes sequenced were classified into multiple variants namely, non-variant of concern/ non-variant of internet (non-VOC/non-VOI (n=140), Eta (n=78), Alpha (n=10), Beta (n=26), Delta (n=171) and Omicron (n=242) (**Figure 1C**).

**Figure 1:**
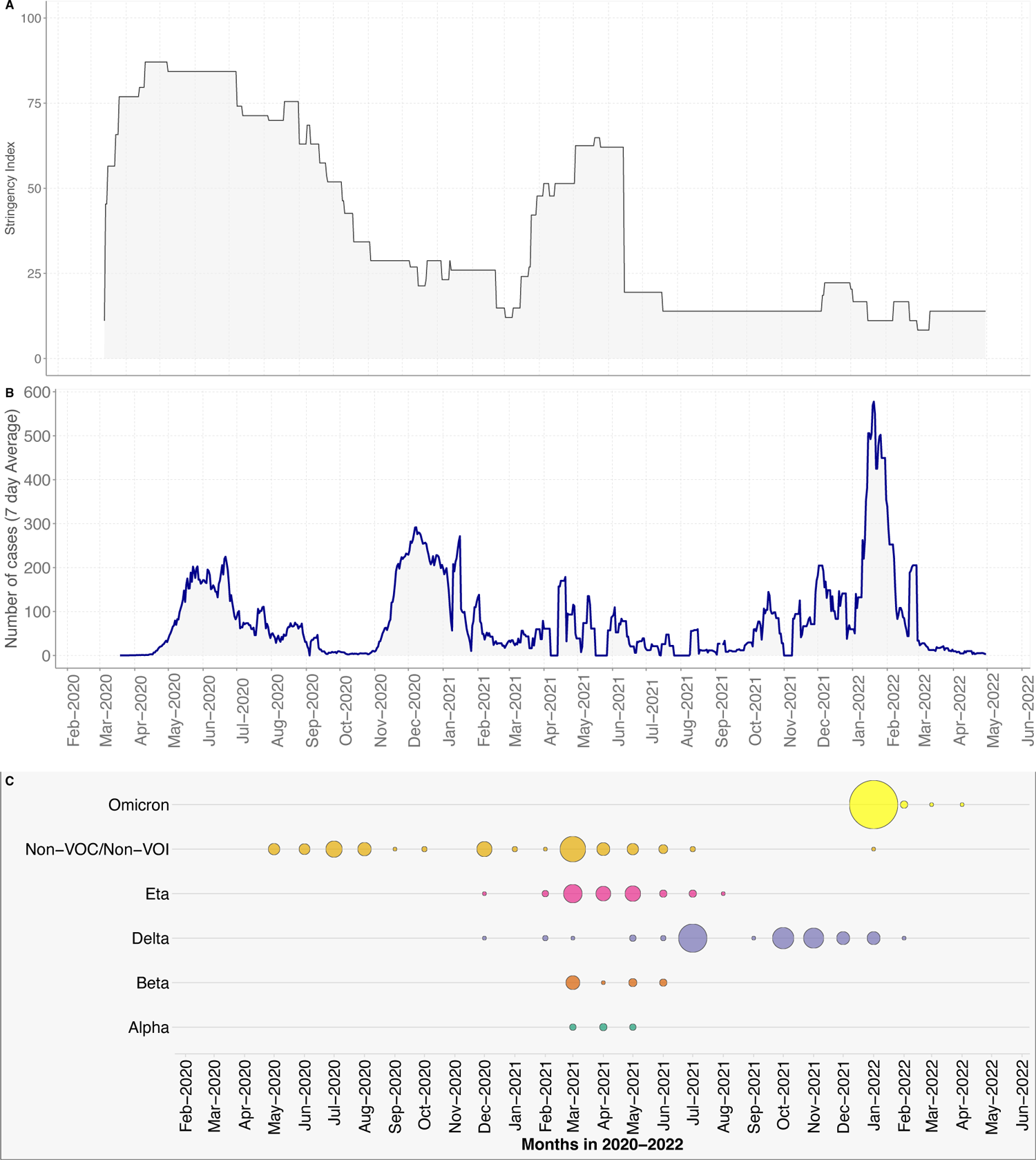
**(A)** Stringency index showing the government intervention levels. **(B)** A weekly rolling average of the daily confirmed SARS-CoV-2 cases in Sudan as populated in https://ourworldindata.org/coronavirus/country/sudan. **(C)** Temporal distribution of detected SARS-CoV-2 variants.

The predominant lineages within the non-VOC/non-VOI group were A.29 (n=43, 30.7%), A (n=42, 30.0%) and B.1 (n=29, 20.7%). These lineages persisted through the first two major waves of SARS-CoV-2 infections in Sudan (Figure 2). The Eta VOI (B.1.525) was first observed in December 2020 and was sustained until August 2021 with the peak cases seen in March 2021 (Figure 2). Alpha and Beta VOC cases were seen to cocirculate between March 2021 and May 2021.

**Figure 2:**
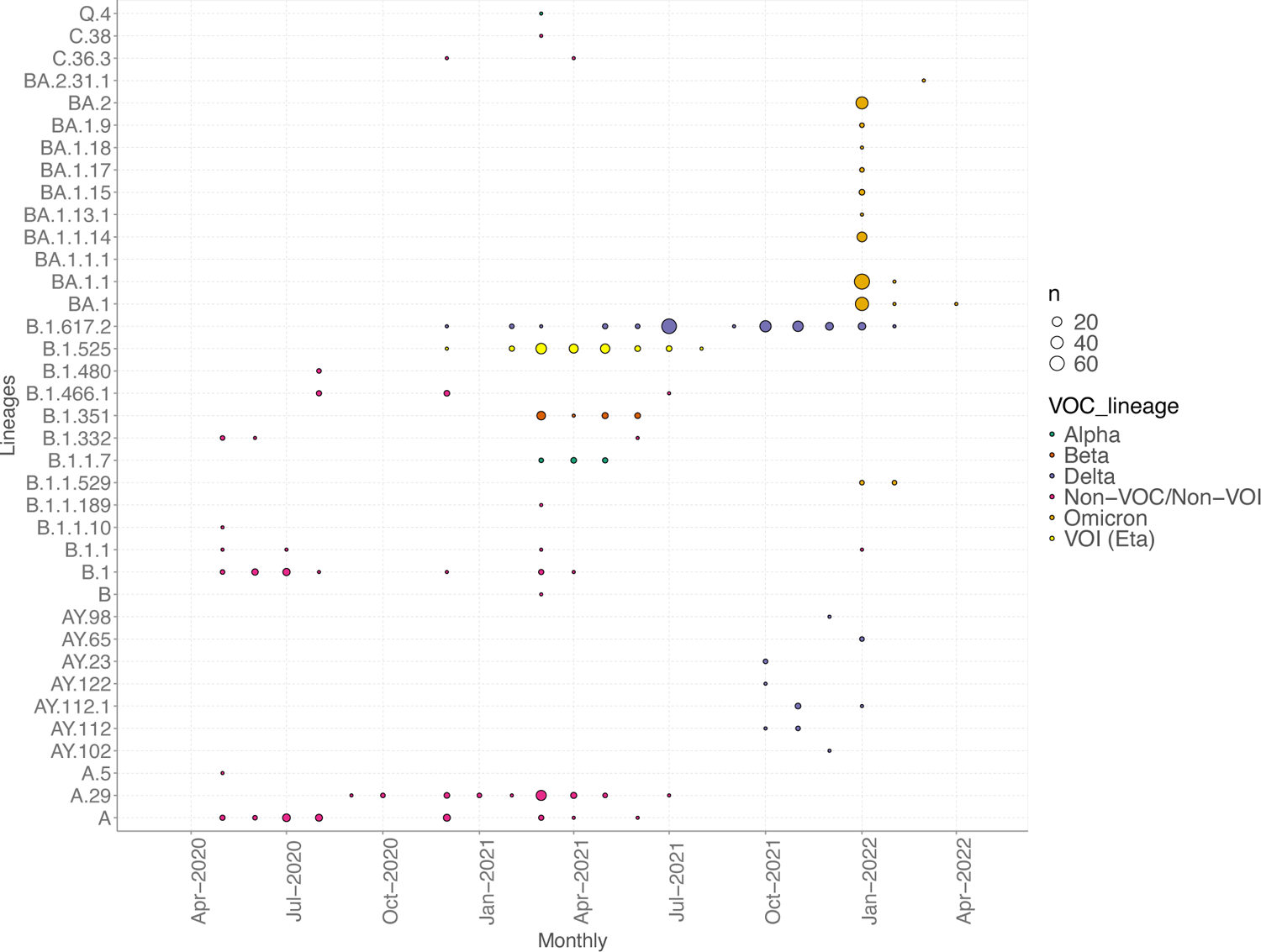
Temporal distribution of detected SARS-CoV-2 lineages in Sudan between May 2020 to April 2022. The colours show the different VOCs/VOI, and the circle sizes indicate the frequency.

Nine Delta VOC lineages were observed between December 2020 and February 2022. The most predominant lineage across this period was B.1.617.2 (n=156, 91.2%). The other Delta VOC lineages were observed between October 2021 and February 2022 (Figure 2). By July 2021, the Delta VOC was the predominant SARS-CoV-2 variant in circulation in Sudan and become dominant by August 2021.

Twelve lineages within the Omicron VOC were observed in between January and April 2022 during the third major wave in Sudan (Figure 2). The most common Omicron lineages were BA.1.1 (n=87, 36.0%), BA.1 (n=60, 24.8%), BA.2 (n=49, 20.2%), and BA.1.1.14 (n=28, 11.6%).

### Genetic diversity of SARS-CoV-2 in Sudan

Multiple clusters and singletons from the Sudan genomes were observed with each of the detected variants. The Sudan genomes were interspersed within the global datasets among all the variants (Figure 3).

**Figure 3:**
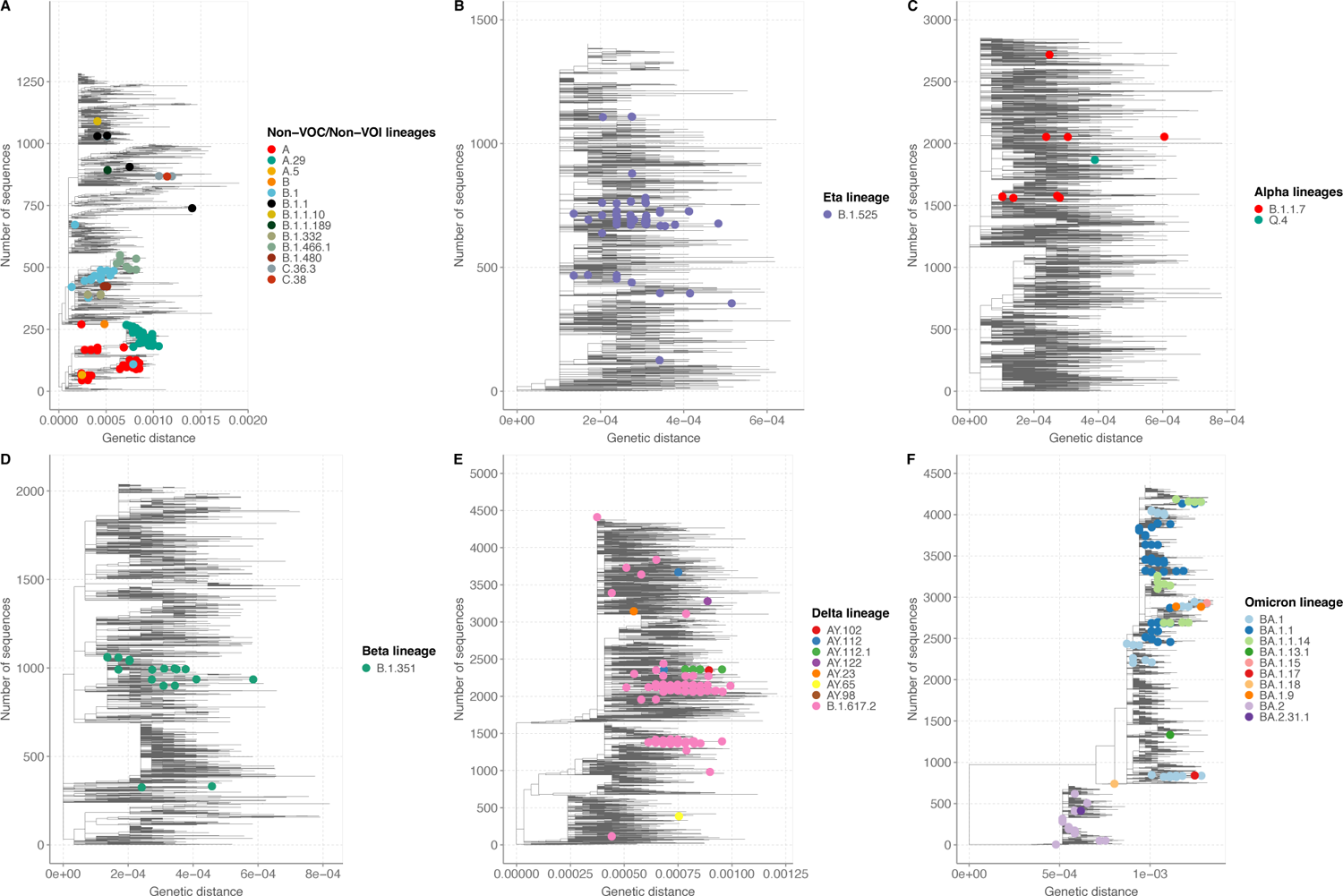
Maximum likelihood phylogenetic trees of SARS-CoV-2 sequences from different lineages **(A)** non-VOC/non-VOI **(B)** Eta VOI **(C)** Alpha VOC **(D)** Beta VOC **(E)** Delta VOC, and **(F)** Omicron VOC from Sudan (coloured dots) and sub-sampled global sequences.

### Importations of SARS-CoV-2 variants in Sudan

Virus importation events into Sudan were estimated from the time-resolved phylogenetic trees. A total of 144 virus introduction events were estimated for all the variants from different parts of the globe. The introduction events for each variant were as follows: non-VOC/non-VOI (n=13), Alpha (n=7), Beta (n=11), Delta (n=24), Eta (n=13) and Omicron (n=76) (**Supplementary** figure 1).

## DISCUSSION

This study describes the genomic epidemiology of SARS-CoV-2 in Sudan between September 2020 and January 2022. Over this period, 37 lineages which were either non-VOCs/non-VOIs Alpha VOC, Eta VOI, Beta VOC, Delta VOC or Omicron VOC were in circulation in Sudan. These SARS-CoV-2 variants sustained the cases throughout 2021 until the third major wave driven by Omicron VOC in January 2022. The third wave coincided with both the relaxation of the stringency measures in Sudan and the introduction of Omicron, with its increased transmissibility and infectivity compared to other VOCs such as Delta and Beta [16], [17].

Lineage A.29 was first detected in Gambia in August 2020 and then spread with minimal infectivity and severity globally [18]. The lineage was first detected in Sudan in September 2020 and was sustained until February 2021 based on additional sequences deposited on GISAID [19]. Alpha VOC, Eta VOI, Beta VOC, and Delta VOC were detected and were in circulation between February 2021 and July 2021. However, no major waves were observed in Sudan coincidented with the introduction of these variants, which contrasts the association of these variants with surges of cases seen in other countries within Africa [6]. The common Omicron lineages detected in cocirculation in Sudan were BA.1.1 (30.7%), BA.1 (27.3%) and BA.2 (23.9%) and the frequency of these variants increased rapidly across the weeks in January 2022. Similar findings in transition of lineages and these omicron subvariants causing surges in cases have been reported elsewhere [20]–[23].

Our phylogenetic analysis showed the sequences from Sudan were interspersed within the global tree, suggesting the frequent global spread of the detected variants due to the movement of people across different countries. Multiple introductions of the Eta VOI, Beta VOC and Omicron VOC were observed into Sudan, mainly from Europe and Africa. Import/export events of the virus within Africa and across Europe have been previously described [6], [8]. Interestingly, only one Delta lineage B.1.617.2 was identified in Sudan and the phylogeny suggests a single introduction event that led further spread of Delta in Sudan through local transmission. However, this conclusion may be influenced by the limited sampling, especially lack of sampling between August and December 2021 following the replacement of the Delta. Our study had some limitations. First, the samples used in this analysis are only from two major cities: Khartoum; and Port Sudan, and we do not have detailed epidemiological data.

Additionally, there is a lack of continuous sampling over time, making it difficult to draw solid conclusions on the complete diversity of SARS-CoV-2 in Sudan during the pandemic.

In conclusion, this study reveals the genetic diversity of SARS-CoV-2 variants in circulation in after multiple introductions of these variants in the country. These findings show that continuous genomic surveillance is key in monitoring and understanding the introduction and spread of SARS-CoV-2 variants within countries as it helps in upholding measures public health measures that can mitigate the spread of the virus.

## Data Availability

The Sudan daily case data for the study period was obtained and is freely available from the Our World in Data database. All genome sequences and associated metadata in this dataset are published in GISAID’s EpiCoV database https://doi.org/10.55876/gis8.231110pw.

The epidemiological data is available on the Virus Epidemiology and Control (VEC) Dataverse (DOI: https://doi.org/10.7910/DVN/782VRG). Data are available under the terms of the <u>Creative Commons Attribution 4.0 International license</u> (CC-BY 4.0).

## ACKNOWLEDGMENTS

We thank the Sudan Ministry of Health, Public Health Laboratory, Africa-CDC for facilitating sharing of sequenced SARS-CoV-2 samples. We would like to thank all the laboratories that have shared SARS-CoV-2 sequence data in GISAID that we included as comparison data in our analysis.

