## SUPPLEMENTRAY TABLE 1 AND SUPPLEMENTARY FIGURE 1 for "Genomic epidemiology of SARS-CoV-2 in Sudan: A retrospective analysis 2020-2022"

**Supplementary table 1: Demographic characteristics of SARS-CoV-2 sequenced samples from Sudan.**

|  | **Omicron (n=242)** | **Delta (n=171)** | **Non-VOC/Non-VOI (N=n=140)** | **VOI (Eta) (n=78)** | **Beta (n=26)** | **Alpha (n=10)** | **Total (n=667)** |
| --- | --- | --- | --- | --- | --- | --- | --- |
| **Location** |  |  |  |  |  |  |  |
| Khartoum | 186 (76.9%) | 86 (50.3%) | 54 (38.6%) | 39 (50.0%) | 23 (88.5%) | 2 (20.0%) | 390 (58.5%) |
| Kassala | 46 (19.0%) | 21 (12.3%) | 70 (50.0%) | 33 (42.3%) | 2 (7.7%) | 8 (80.0%) | 180 (27.0%) |
| Portsudan | 0 (0.0%) | 59 (34.5%) | 5 (3.6%) | 2 (2.6%) | 0 (0.0%) | 0 (0.0%) | 66 (9.9%) |
| Western Kordofan | 0 (0.0%) | 0 (0.0%) | 1 (0.7%) | 2 (2.6%) | 0 (0.0%) | 0 (0.0%) | 3 (0.4%) |
| Jabal Awlya | 1 (0.4%) | 2 (1.2%) | 0 (0.0%) | 0 (0.0%) | 0 (0.0%) | 0 (0.0%) | 3 (0.4%) |
| East nile | 0 (0.0%) | 1 (0.6%) | 0 (0.0%) | 0 (0.0%) | 0 (0.0%) | 0 (0.0%) | 1 (0.1%) |
| Eldoum | 1 (0.4%) | 0 (0.0%) | 0 (0.0%) | 0 (0.0%) | 0 (0.0%) | 0 (0.0%) | 1 (0.1%) |
| Elobyed | 1 (0.4%) | 0 (0.0%) | 0 (0.0%) | 0 (0.0%) | 0 (0.0%) | 0 (0.0%) | 1 (0.1%) |
| Sinar | 0 (0.0%) | 1 (0.6%) | 0 (0.0%) | 0 (0.0%) | 0 (0.0%) | 0 (0.0%) | 1 (0.1%) |
| Western Nile | 0 (0.0%) | 0 (0.0%) | 1 (0.7%) | 0 (0.0%) | 0 (0.0%) | 0 (0.0%) | 1 (0.1%) |
| Whitenile | 0 (0.0%) | 1 (0.6%) | 0 (0.0%) | 0 (0.0%) | 0 (0.0%) | 0 (0.0%) | 1 (0.1%) |
| No data | 7 (2.9%) | 0 (0.0%) | 9 (6.4%) | 2 (2.6%) | 1 (3.8%) | 0 (0.0%) | 19 (2.8%) |
| **Gender** |  |  |  |  |  |  |  |
| Female | 85 (35.1%) | 82 (48.0%) | 53 (37.9%) | 33 (42.3%) | 11 (42.3%) | 3 (30.0%) | 267 (40.0%) |
| Male | 150 (62.0%) | 89 (52.0%) | 78 (55.7%) | 43 (55.1%) | 14 (53.8%) | 7 (70.0%) | 381 (57.1%) |
| No data | 7 (2.9%) | 0 (0.0%) | 9 (6.4%) | 2 (2.6%) | 1 (3.8%) | 0 (0.0%) | 19 (2.8%) |
| **Age group (years)** |  |  |  |  |  |  |  |
| 0 - 9 | 13 (5.4%) | 0 (0.0%) | 1 (0.7%) | 0 (0.0%) | 0 (0.0%) | 0 (0.0%) | 14 (2.1%) |
| 10 - 19 | 17 (7.0%) | 14 (8.2%) | 5 (3.6%) | 6 (7.7%) | 4 (15.4%) | 0 (0.0%) | 46 (6.9%) |
| 20 - 29 | 32 (13.2%) | 37 (21.6%) | 22 (15.7%) | 9 (11.5%) | 7 (26.9%) | 1 (10.0%) | 108 (16.2%) |
| 30 - 39 | 33 (13.6%) | 33 (19.3%) | 26 (18.6%) | 17 (21.8%) | 5 (19.2%) | 1 (10.0%) | 115 (17.2%) |
| 40 - 49 | 31 (12.8%) | 29 (17.0%) | 14 (10.0%) | 12 (15.4%) | 2 (7.7%) | 1 (10.0%) | 89 (13.3%) |
| 50 - 59 | 28 (11.6%) | 25 (14.6%) | 18 (12.9%) | 10 (12.8%) | 4 (15.4%) | 0 (0.0%) | 85 (12.7%) |
| ≥60 | 30 (12.4%) | 30 (17.5%) | 36 (25.7%) | 21 (26.9%) | 3 (11.5%) | 7 (70.0%) | 127 (19.0%) |
| No data | 58 (24.0%) | 3 (1.8%) | 18 (12.9%) | 3 (3.8%) | 1 (3.8%) | 0 (0.0%) | 83 (12.4%) |
| **Clinical Presentation** |  |  |  |  |  |  |  |
| Symptomatic | 95 (39.3%) | 85 (49.7%) | 9 (6.4%) | 32 (41.0%) | 11 (42.3%) | 5 (50.0%) | 237 (35.5%) |
| Asymptomatic | 90 (37.2%) | 82 (48.0%) | 1 (0.7%) | 6 (7.7%) | 0 (0.0%) | 0 (0.0%) | 179 (26.8%) |
| No data | 57 (23.6%) | 4 (2.3%) | 130 (92.9%) | 40 (51.3%) | 15 (57.7%) | 5 (50.0%) | 251 (37.6%) |
| **Travel History** |  |  |  |  |  |  |  |
| Yes | 0 (0.0%) | 1 (0.6%) | 0 (0.0%) | 2 (2.6%) | 1 (3.8%) | 0 (0.0%) | 4 (0.6%) |
| No | 185 (76.4%) | 108 (63.2%) | 9 (6.4%) | 34 (43.6%) | 10 (38.5%) | 5 (50.0%) | 351 (52.6%) |
| No data | 57 (23.6%) | 62 (36.3%) | 131 (93.6%) | 42 (53.8%) | 15 (57.7%) | 5 (50.0%) | 312 (46.8%) |
| **Reason for testing** |  |  |  |  |  |  |  |
| Presented to health facility | 5 (2.1%) | 0 (0.0%) | 0 (0.0%) | 0 (0.0%) | 0 (0.0%) | 0 (0.0%) | 5 (0.7%) |
| Contact with confirmed cases | 6 (2.5%) | 1 (0.6%) | 0 (0.0%) | 0 (0.0%) | 0 (0.0%) | 0 (0.0%) | 7 (1.0%) |
| No data | 231 (95.5%) | 170 (99.4%) | 140 (100.0%) | 78 (100.0%) | 26 (100.0%) | 10 (100.0%) | 655 (98.2%) |
| **Vaccination Status** |  |  |  |  |  |  |  |
| Fully vaccinated | 24 (9.9%) | 0 (0.0%) | 0 (0.0%) | 0 (0.0%) | 0 (0.0%) | 0 (0.0%) | 24 (3.6%) |
| Not vaccinated | 95 (39.3%) | 2 (1.2%) | 1 (0.7%) | 0 (0.0%) | 0 (0.0%) | 0 (0.0%) | 98 (14.7%) |
| No data | 123 (40.8%) | 169 (98.8%) | 139 (99.3%) | 78 (100.0%) | 26 (100.0%) | 10 (100.0%) | 545 (81.7%) |

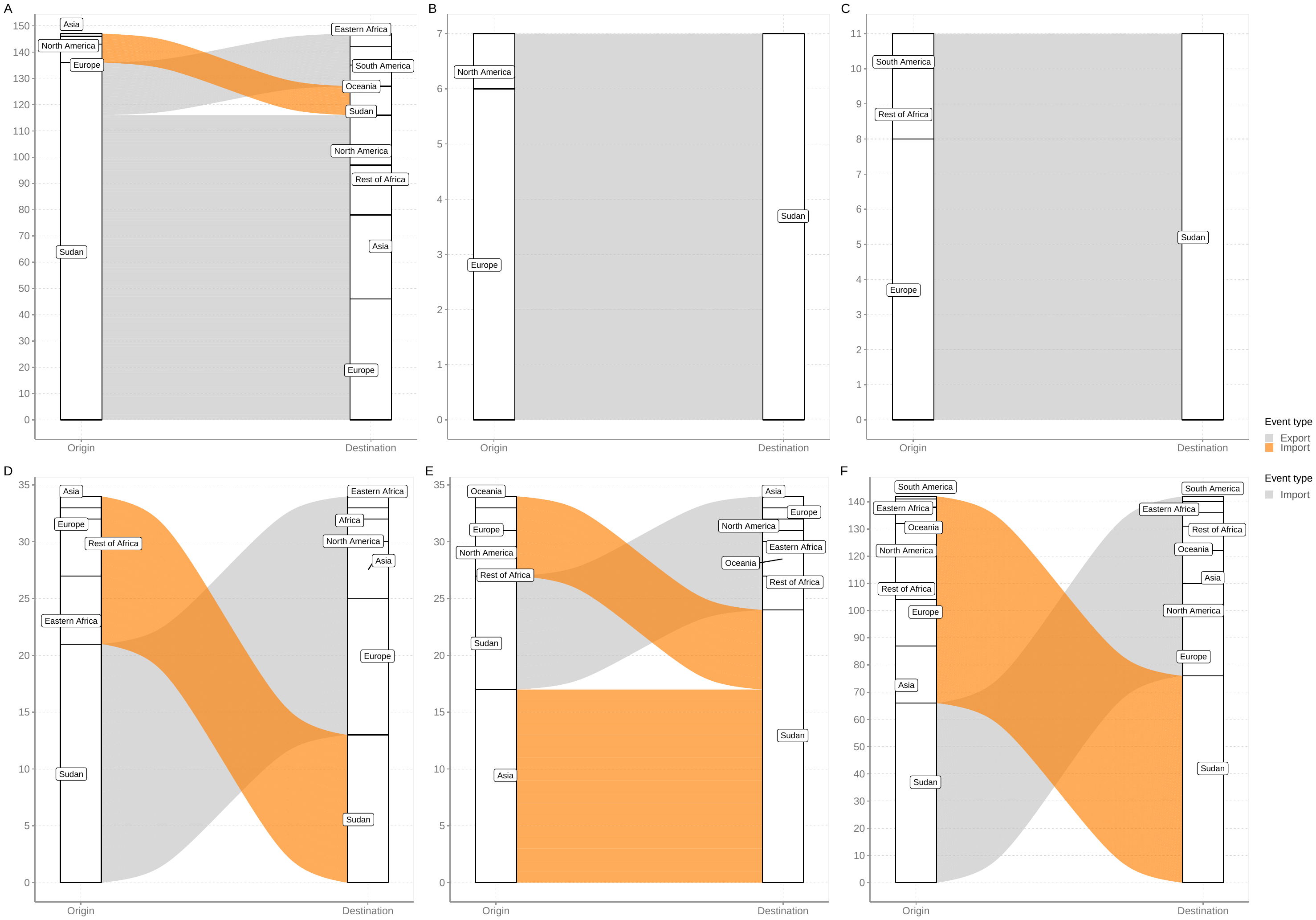

**Supplementary figure1:** Mugration plots showing SARS-CoV-2 import/export events in Sudan from different regions globally. (A) non-VOC/non-VOI (B)Alpha VOC, (C) Beta VOC, (D) Eta VOI (E) Delta VOC and (F) Omicron VOC
